# Birthweight by gestational age reference centile charts for Greek neonates

**DOI:** 10.1101/2020.10.04.20204040

**Authors:** A Tsagkari, K Pateras, D Ladopoulou, E Kornarou, N Vlachadis

## Abstract

**Objective:** The development of population based, sex-specific, birthweight for gestational age (GA) first-time reference centile charts for liveborn singletons in Greece.

**Design:** Secondary analysis of national birth registry data

**Participants:** Data of birthweight by GA of all 633201 singleton live births in Greece between 2011 and 2017 were collected from the Hellenic Statistical Authority (ELSTAT).

**Main outcome measures:** After excluding implausible birthweights, we estimated gender specific birthweight centiles for every gestational week from 22^nd^ to 40^th^+ using the Lambda-Mu-Sigma (LMS) method via the GAMLSS package in R. Small (10^th^ centile) and large-for GA (90^th^ centile) cut-offs in certain gestational weeks were compared to previously published charts.

**Results:** More than 90% of the 621043 neonates were born at term (>37 weeks of GA). The mean birthweight for boys and girls at 40+ weeks of GA were 3473 grams and 3327 grams respectively. Most neonates were born at 38 weeks of GA with a mean birth-weight of 3097 (97655 girls) and 3237 (104722 boys) grams. The proposed national centiles identified more or less deviant percentages of small and large for GA neonates in comparison to other (inter)national centiles.

**Conclusions:** The new centile charts provide important information on the contemporary distribution of birthweight for gestational age in Greece. They may assist physicians to classify high-risk neonates at birth based on national population data. Accurate identification of those in need of special care will help to prevent possible adverse sequelae in the perinatal period and beyond.

## INTRODUCTION

Neonates that largely deviate from the appropriate birthweight for their gestational age (GA) are prone to increased mortality and morbidity in the perinatal and neonatal period and they also show increased incidence of health problems in adult life, namely obesity, metabolic syndrome, type II diabetes, neurodevelopmental disorders (1–5).

Physicians estimate and monitor intrauterine growth based on measurement of weight (and/or other fetal/neonatal parameters) for each GA. Data of weight by GA are depicted on centile charts and their corresponding centile curves. Currently three different types of growth charts are used, depending on the study design and population selection; prescriptive (normative), descriptive (reference) and customized charts.

Prescriptive (normative) charts usually result from prospective, longitudinal studies on a selected low-risk pregnancy population and they are more often proposed for monitoring fetal growth. These charts show how fetuses should grow, when there are no inhibitive factors present for their intrauterine growth (6). Descriptive (reference) charts usually result from cross-sectional studies on an unselected population and they show how fetuses have grown within a specific population and within a specific time period in the past. They were first employed for accurate classification of high risk neonates, including those affected by intrauterine growth restriction (7). These charts often assist physicians for the assessment of individual infants’ growth at birth, compared to infants of the local population. Descriptive charts are also used by public health researchers as they provide epidemiological information on the distributions of birthweight by GA within the specific population and within a specific time period. These charts often assist public health policy makers to better allocate scarce healthcare resources within the perinatal period (8). Last but not least, the customized growth charts (Gestation Related Optimal Weight-GROW-charts) (9) constitute a third type of charts lying between the two aforementioned. This latter type, borrows features of both prescriptive and descriptive charts. Its creation is based on a statistical process that estimates the optimal weight of the fetus for each GA accounting for certain maternal and fetal characteristics (10). To add further complexity, a generic reference for fetal weight and birthweight percentiles, claimed to be easily adapted for local populations has been proposed by Mikolajczyk et al (11). Up until now however, there seems to be no global consensus about the type that should be used for the assessment of fetal growth (12).

Most developed countries have created birthweight for GA reference charts that are based on cross sectional studies using registry data from their official population records. To the best of our knowledge, in Greece, such a national reference chart does not yet exist. In practice physicians in Greek neonatal units are utilizing the revised 2013 Fenton preterm growth charts to assign size, -small, appropriate and large- for GA at birth (13), even though, the percentage rate for each size category with these charts has been shown to differ compared to local population data and accordingly may not be representative of local neonatal populations (14).

The main aim of this study is to create first-time national birthweight by GA percentiles for all male and female neonates born in Greece over a 7-year period between 2011 and 2017. A comparison with previously published centile charts is also presented.

## METHODS

Data of all singleton live born neonates from 20 to 40+ weeks GA in Greece between 2011 and 2017, were obtained from the Hellenic statistical authority (ELSTAT). The Hellenic statistical authority (ELSTAT) until 2016 added up neonates with GAs of over 40 weeks to those with 40 weeks of GA. This is why the last gestational week in the present study was defined as 40+. Neonatal data of 2017 were accordingly adapted (Neonates with 41 and 42 gestational weeks were added up to the number of infants with 40 weeks of GA). Neonates with missing birthweight and/or GA were excluded from this study. In addition, outliers (neonates with implausible birthweight for GA) (15), were excluded by using 2 distinct statistical methods (Supplementary file) depending on the GA (16,17,18).

Birthweight centiles were estimated with the LMS method via GAMLSS package, while assuming that the birthweight has a Box-Cox and Green distribution as recommended by WHO (6). The LMS approach transforms skewed to approximate normal distributions. To perform that, the LMS function determines the optimal number of effective degrees of freedom and then it estimates the L, M and S parameters; namely the Box-Cox power (L), the median (M) and the coefficient of variation (S) and determines an optimal number of effective degrees of freedom. GAs were transformed to the log scale and the penalized splines (‘pb’) function was applied. This P-splines smoother function utilizes a maximum likelihood estimation to select effective degrees of freedom that comprise the best model. Then, the smoothed values based on L, M and S were utilized to transform the observed distribution of birthweight to a standard normal distribution. The LMS method via GAMLSS (15) was implemented in R version 3.5.3 (19,20).

Subsequently, we performed a comparison that was based on the 10th and 90th birthweight centile values at GA 24, 28, 32 and 36 weeks of the present study versus 3 international normative (WHO fetal, Intergrowth-21st Fetal and Intergrowth-21st Neonatal (6,21,22), 6 national (population) reference curves of European and non-European countries (23-28) and the revised 2013 Fenton growth charts for preterm infants (13).

## RESULTS

633201 singleton neonates were born in Greece between 2011 and 2017 (male/female ratio 1,06:1) with GAs of 20 to 40+ weeks. Out of the 633201 neonates, a) 6273 (0,99%) neonates with missing birthweight, b) 3134 (0,49%) with missing both birthweight and GA and c) 1834 (0,29%) with missing GA were excluded. In addition, 4 neonates with less than 22 weeks of GA and 913 (0,14%) neonates whose birthweight was considered implausible, were excluded as well. The resulting study population consisted of 621043 (320397 male and 300646 female) live born singletons (98,08%) with 22 to 40+ weeks of GA. In Table 1, the distribution of neonates by birthweight, GA and gender is presented. As expected, the majority of neonates (∼90%), irrespectively of gender, are born with GAs ≥ 37 weeks (term births), while the most frequent GA at birth is the 38^th^ week for both male and female neonates. Table 1 also presents 9 distributional centiles of birthweight by GA, as well as the mean birthweight and standard deviation for each GA. Among GAs, the minimum recorded mean birthweight was 607 and 523 grams and the maximum recorded mean birthweight was 3473 and 3327 grams, for male and female neonates respectively.

**Table 1.** Birthweight (in grams) by gestational age centiles– Males and Females born in Greece (2011-2017)

|  | Males (Total Number: 320.397) |  |  |  |  |  |  |  |  |  |  |  |  | Females (Total Number: 300.646) |  |  |  |  |  |  |  |  |  |  |  |
| --- | --- | --- | --- | --- | --- | --- | --- | --- | --- | --- | --- | --- | --- | --- | --- | --- | --- | --- | --- | --- | --- | --- | --- | --- | --- |
| Gestational age | N<sup>1</sup> | Mean | SD | 3rd | 5th | 10th | 25th | 50th | 75th | 90th | 95th | 97th |  | N | Mean | SD | 3rd | 5th | 10th | 25th | 50th | 75th | 90th | 95th | 97th |
| 22 | 21 | 607 | 119 | 401 | 427 | 463 | 517 | 573 | 631 | 692 | 735 | 767 |  | 20 | 523 | 89 | 383 | 403 | 431 | 475 | 523 | 571 | 617 | 646 | 667 |
| 23 | 41 | 633 | 86 | 443 | 471 | 511 | 573 | 637 | 703 | 768 | 813 | 846 |  | 39 | 589 | 88 | 430 | 451 | 482 | 532 | 585 | 638 | 688 | 720 | 741 |
| 24 | 84 | 708 | 118 | 487 | 520 | 566 | 638 | 713 | 789 | 862 | 912 | 946 |  | 66 | 664 | 84 | 478 | 502 | 537 | 594 | 655 | 716 | 772 | 808 | 831 |
| 25 | 126 | 796 | 131 | 537 | 576 | 631 | 717 | 807 | 897 | 982 | 1038 | 1076 |  | 101 | 718 | 103 | 521 | 550 | 594 | 663 | 738 | 812 | 880 | 921 | 949 |
| 26 | 190 | 916 | 186 | 592 | 639 | 707 | 812 | 921 | 1029 | 1130 | 1194 | 1238 |  | 142 | 841 | 152 | 558 | 596 | 652 | 742 | 838 | 933 | 1019 | 1071 | 1105 |
| 27 | 218 | 1.069 | 179 | 649 | 707 | 790 | 918 | 1050 | 1178 | 1296 | 1370 | 1420 |  | 186 | 949 | 197 | 592 | 642 | 715 | 832 | 956 | 1078 | 1187 | 1253 | 1296 |
| 28 | 313 | 1.170 | 256 | 707 | 776 | 876 | 1028 | 1184 | 1335 | 1471 | 1555 | 1610 |  | 249 | 1.074 | 233 | 635 | 697 | 789 | 935 | 1090 | 1241 | 1376 | 1457 | 1510 |
| 29 | 322 | 1.304 | 267 | 778 | 859 | 974 | 1150 | 1330 | 1502 | 1655 | 1748 | 1809 |  | 261 | 1.235 | 275 | 700 | 773 | 881 | 1055 | 1240 | 1419 | 1579 | 1674 | 1736 |
| 30 | 512 | 1.485 | 303 | 877 | 967 | 1097 | 1295 | 1498 | 1690 | 1860 | 1962 | 2029 |  | 421 | 1.388 | 322 | 799 | 880 | 1001 | 1195 | 1403 | 1604 | 1783 | 1889 | 1958 |
| 31 | 555 | 1.660 | 344 | 1011 | 1108 | 1249 | 1466 | 1688 | 1900 | 2087 | 2199 | 2272 |  | 522 | 1.565 | 321 | 938 | 1024 | 1152 | 1359 | 1579 | 1795 | 1986 | 2100 | 2175 |
| 32 | 1029 | 1.889 | 355 | 1179 | 1281 | 1429 | 1658 | 1895 | 2122 | 2324 | 2446 | 2527 |  | 854 | 1.759 | 352 | 1110 | 1199 | 1332 | 1546 | 1776 | 2001 | 2203 | 2324 | 2403 |
| 33 | 1477 | 2.094 | 372 | 1369 | 1473 | 1625 | 1862 | 2110 | 2350 | 2568 | 2701 | 2789 |  | 1260 | 1.990 | 362 | 1308 | 1399 | 1536 | 1756 | 1992 | 2226 | 2438 | 2567 | 2653 |
| 34 | 2806 | 2.354 | 430 | 1580 | 1686 | 1841 | 2083 | 2338 | 2590 | 2824 | 2972 | 3071 |  | 2461 | 2.252 | 422 | 1507 | 1604 | 1749 | 1978 | 2224 | 2470 | 2701 | 2846 | 2945 |
| 35 | 5167 | 2.589 | 408 | 1824 | 1931 | 2088 | 2331 | 2588 | 2847 | 3095 | 3257 | 3369 |  | 4580 | 2.478 | 415 | 1721 | 1826 | 1978 | 2217 | 2471 | 2731 | 2983 | 3150 | 3266 |
| 36 | 14025 | 2.856 | 409 | 2099 | 2205 | 2357 | 2595 | 2849 | 3107 | 3360 | 3527 | 3645 |  | 12423 | 2.728 | 397 | 2000 | 2098 | 2242 | 2471 | 2718 | 2974 | 3225 | 3391 | 3508 |
| 37 | 38479 | 3.077 | 393 | 2353 | 2451 | 2596 | 2824 | 3070 | 3322 | 3568 | 3731 | 3844 |  | 34783 | 2.941 | 378 | 2254 | 2345 | 2480 | 2695 | 2930 | 3174 | 3413 | 3571 | 3682 |
| 38 | 104702 | 3.237 | 384 | 2536 | 2629 | 2768 | 2988 | 3229 | 3476 | 3716 | 3873 | 3982 |  | 97655 | 3.097 | 367 | 2433 | 2520 | 2650 | 2859 | 3087 | 3324 | 3557 | 3709 | 3816 |
| 39 | 78690 | 3.365 | 384 | 2665 | 2757 | 2894 | 3114 | 3356 | 3605 | 3845 | 4001 | 4108 |  | 75334 | 3.223 | 366 | 2560 | 2646 | 2775 | 2983 | 3213 | 3451 | 3683 | 3833 | 3938 |
| 40+ | 71640 | 3.473 | 404 | 2737 | 2834 | 2978 | 3210 | 3465 | 3726 | 3978 | 4141 | 4253 |  | 69289 | 3.327 | 385 | 2630 | 2721 | 2856 | 3076 | 3317 | 3567 | 3809 | 3967 | 4076 |

<sup>1</sup> N: Number of neonates

The proposed smoothed (corrected) sex-specific reference centile curves for neonates born in Greece are presented in Figures 1 and 2. These curves were produced after correcting the crude (not corrected) curves for implausible birthweights and/or possibly misclassified neonates. Such a misclassification is most prominent in earlier GAs (<30 weeks) as illustrated in the crude reference data and curves that are presented in the supplementary file (Figure 1A and Figure 2A).

**Figure 1.**
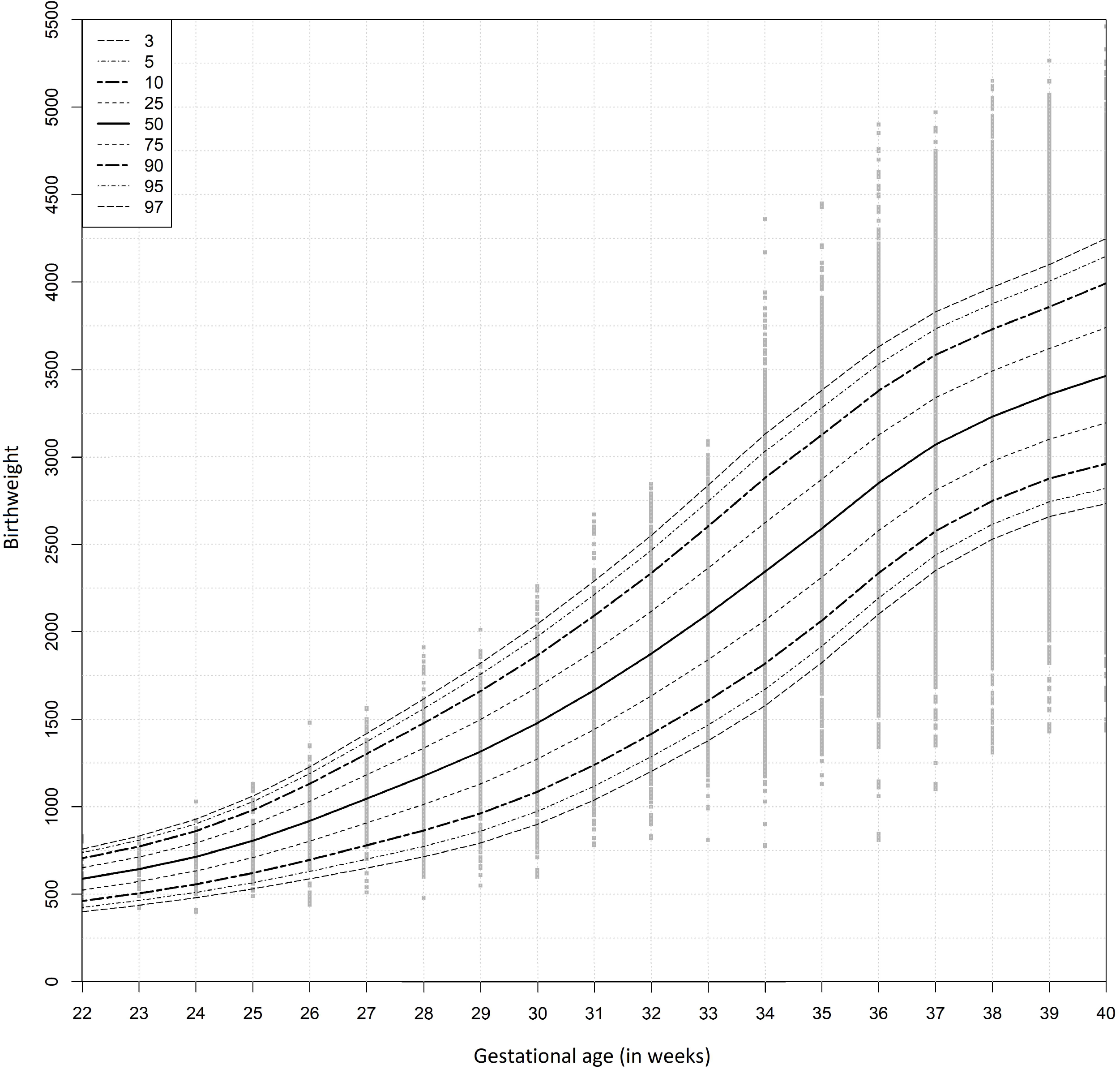
Birthweight (in grams) by gestational age (weeks) smoothed curves - Males born in Greece between 2011 and 2017.

**Figure 2.**
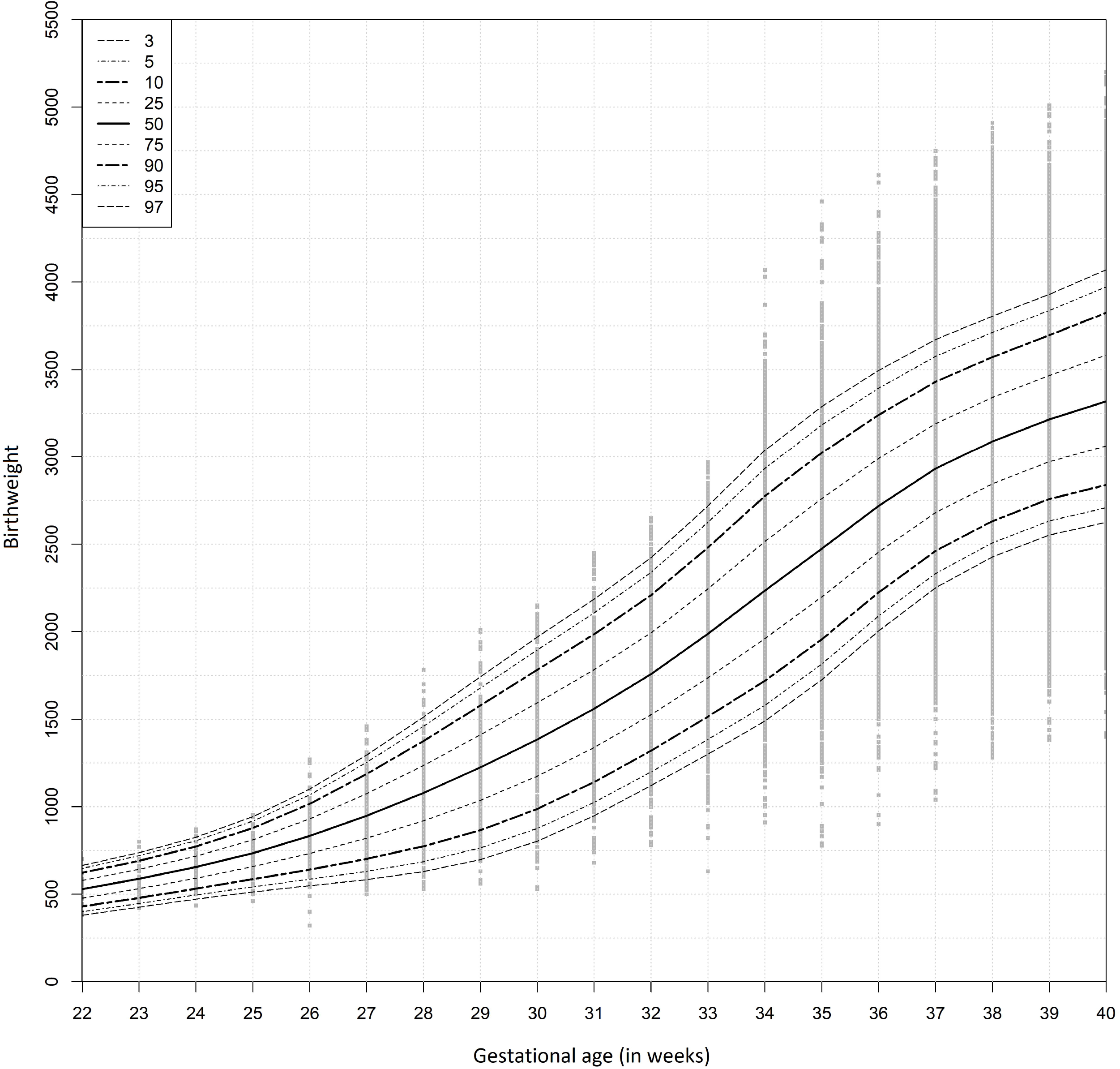
Birthweight (in grams) by gestational age (weeks) smoothed curves Females born in Greece between 2011 and 2017.

Table 2 presents the absolute differences in birthweight and the relative percentual differences at the 10th and 90th percentiles of birthweight, between previously published references and the proposed reference, at the 24^th^, 28^th^, 32^nd^ and 36^th^ gestational week. For example, at the 36^th^ week of GA, the proposed reference identifies 9,6% of male neonates as small-for GA. If the Fenton curves are used in the Greek population, 2,2% fewer male infants will be identified as small-for GA in comparison to the proposed reference. This percentual difference corresponds to almost 308 male infants in the study period, since the number of males born with 36 weeks of GA is 14025 (Table 1). The proposed reference generally identifies more neonates as small-for GA compared to Fenton curves, especially in the very early gestational weeks (24^th^ and 28^th^). The large-for GA rate slightly differs among the proposed and Fenton chart across the four studied gestational weeks in both genders (Table 2). Overall, among the national centiles, the Canadian and Italian ones produce percentages that are more in line with the Greek proposed reference centiles at the specific gestational weeks, while the Dutch, Brazilian and USA centiles produce the most deviant percentages from the Greek centiles, especially regarding large-for GA neonates (Table 2). Finally, the proposed reference identifies significantly less small-for GA neonates compared to the WHO and Intergrowth 21^st^ fetal centiles up to the 32^nd^ week (Table 2).

**Table 2.** Absolute differences in birthweight and relative percentual differences at the 10^th^ and 90^th^ percentiles of birth weight between the previously published references and the proposed reference^2^ at certain gestational weeks.

| Gestational Age | 24 |  | 28 |  | 32 |  | 36 |  | 24 |  | 28 |  | 32 |  | 36 |  |
| --- | --- | --- | --- | --- | --- | --- | --- | --- | --- | --- | --- | --- | --- | --- | --- | --- |
| Small for gestational age<br>SGA (10th centile) | Male | Female | Male | Female | Male | Female | Male | Female | Male | Female | Male | Female | Male | Female | Male | Female |
| Proposed Reference (PR) | 566gr | 537gr | 876gr | 789gr | 1429gr | 1332gr | 2357gr | 2242gr | 14,3% | 6,1% | 13,7% | 12,4% | 11,1% | 13,7% | 9,6% | 10,2% |
| Previously Published (PP) | PP-PR (Δgr) |  | PP-PR (Δgr) |  | PP-PR (Δgr) |  | PP-PR (Δgr) |  | PP-PR (Δ%) |  | PP-PR (Δ%) |  | PP-PR (Δ%) |  | PP-PR (Δ%) |  |
| FENTON | -19 | -22 | -81 | -51 | -13 | 5 | -60 | -45 | -6,0 | -1,5 | -5,1 | -3,6 | -0,5 | 0,1 | -2,2 | -1,9 |
| WHO [13] | 26 | 30 | 179 | 224 | 248 | 282 | 47 | 72 | 3,6 | 4,5 | 16,3 | 23,7 | 13,5 | 18,6 | 2,5 | 3,6 |
| INTERGROWTH 21 [14] | 36 | 65 | 75 | 162 | 44 | 141 | -213 | -98 | 6,0 | 16,7 | 5,4 | 16,5 | 1,7 | 7,5 | -5,7 | -3,8 |
| INTERGROWTH 21 [15] | -- | -- | -- | -- | -- | -- | -177 | -102 | -- | -- | -- | -- | -- | -- | -5,2 | -4,1 |
| Canadian curves [17] | -19 | -24 | -23 | 13 | 15 | 14 | -36 | -15 | -6,0 | -1,5 | -1,6 | 3,2 | 0,9 | 0,9 | -1,2 | -0,7 |
| Scottish curves [21] | -126 | -148 | -110 | -93 | -47 | -69 | -153 | -143 | -10,7 | -6,1 | -6,1 | -6,0 | -1,3 | -4,6 | -4,4 | -5,4 |
| Dutch curves [18] | -- | -- | -146 | -169 | -200 | -229 | -226 | -214 | -- | -- | -8,9 | -9,2 | -7,1 | -11,0 | -5,9 | -6,8 |
| Brazilian curves [19] | -49 | -37 | -106 | -54 | -102 | -51 | -152 | -82 | -8,3 | -3,0 | -6,1 | -3,6 | -4,2 | -3,9 | -4,4 | -3,3 |
| Italian curves [16] | 7 | 23 | -63 | -106 | -106 | -115 | -137 | -145 | 0,0 | 3,0 | -3,8 | -6,4 | -4,3 | -7,4 | -4,2 | -5,4 |
| USA curves [20] | -68 | -39 | -78 | 9 | 66 | 163 | -3 | 112 | -10,7 | -4,5 | -5,1 | 0,8 | 2,3 | 8,7 | 0,0 | 6,2 |
| Large for gestational age LGA (90th centile) |  |  |  |  |  |  |  |  |  |  |  |  |  |  |  |  |
| Proposed Reference (PR) | 862gr | 772gr | 1471gr | 1376gr | 2324gr | 2203gr | 3360gr | 3225gr | 8,3% | 7,6% | 10,9% | 6,4% | 9,6% | 10,9% | 10% | 10,1% |
| Previously Published (PP) | PP-PR (Δgr) |  | PP-PR (Δgr) |  | PP-PR (Δgr) |  | PP-PR (Δgr) |  | PP-PR (Δ%) |  | PP-PR (Δ%) |  | PP-PR (Δ%) |  | PP-PR (Δ%) |  |
| FENTON | -17 | 16 | -38 | -24 | -20 | 15 | 30 | 71 | 0,0 | 0,0 | 1,9 | 1,6 | 0,8 | -4,9 | -0,9 | -2,4 |
| WHO [13] | -77 | -32 | -72 | -53 | -100 | -76 | -175 | -126 | 15,5 | 6,1 | 5,8 | 4,0 | 6,5 | 4,4 | 9,1 | 6,9 |
| INTERGROWTH 21 [14] | -111 | -21 | -195 | -100 | -235 | -114 | -271 | -136 | 26,2 | 3,0 | 21,4 | 10,0 | 17,5 | 6,6 | 17,0 | 7,3 |
| INTERGROWTH 21 [15] | -- | -- | -- | -- | -- | -- | -110 | -105 | -- | -- | -- | -- | -- | -- | 4,7 | 4,5 |
| Canadian curves [17] | -18 | 18 | -26 | 14 | -5 | 39 | 51 | 82 | 0,0 | 0,0 | 1,9 | -0,4 | 0,6 | -1,6 | -1,7 | -2,9 |
| Scottish curves [21] | -6 | 17 | -71 | -49 | -52 | -40 | -82 | -43 | 0,0 | 0,0 | 4,5 | 4,0 | 2,8 | 2,0 | 3,8 | 1,7 |
| Dutch curves [18] | -- | -- | -160 | -168 | -171 | -142 | -131 | -94 | -- | -- | 15,0 | 22,1 | 11,3 | 7,8 | 6,6 | 4,1 |
| Brazilian curves [19] | 171 | 220 | 78 | 151 | 291 | 391 | 188 | 231 | -8,3 | -7,6 | -3,2 | -3,6 | -7,8 | -10 | -5,1 | -6,1 |
| Italian curves [16] | -15 | 65 | -48 | -36 | -44 | -23 | -97 | -95 | 0,0 | -4,5 | 3,2 | 2,0 | 2,1 | 1,5 | 4,2 | 4,1 |
| USA curves [20] | 115 | 205 | 506 | 601 | 876 | 997 | 308 | 443 | -7,1 | -7,6 | -11 | -6,4 | -9,6 | -11 | -7,3 | -8,7 |

<sup>2</sup> (Positive numbers are shown when the previously published percentiles' respective values are larger than in the proposed one. This positive sign suggests that more neonates would have been identified as e.g. SGA (overestimation) if the older centiles were used in the current Greek population. Negative numbers suggest that less neonates would have been identified as e.g. SGA (underestimation) if the older centiles were used instead of the proposed one in the study population).

## DISCUSSION

In clinical practice, physicians attain useful information by comparing the birthweight of a neonate at a specific GA with the birthweights of neonates of the same GA in the same general population. Such comparisons allow for a more accurate classification of neonates at increased risk for perinatal complications. In addition, the impact of inhibitive or strengthening factors for fetal growth in the specific local population may be associated with this neonatal classification.

?n Greece, there is no recommendation on how to assess growth prenatally and the classification of neonates as small, appropriate and large-for-GA at birth is not made according to national population data (2). Obstetricians use charts based on Hadlock formula (29) or various charts incorporated as default tools in ultrasound programs for estimating and monitoring fetal growth and assessing possible intrauterine growth restriction. Neonatologists and pediatricians in the Greek neonatal units use the 2013 revised Fenton preterm growth charts to assign size at birth, as these charts offer the additional convenience of postnatal preterm infant growth monitoring (13).

However, the use of multiethnic or international charts should not always be considered a panacea, as it may not be representative of local populations and it may lead to misclassifications of high risk neonates (14,29,30,31). In a recent study on intrauterine growth references, the prevalence of small-for GA neonates was shown to differ among 11 European countries (32). When various European reference charts based on the Gardosi model were applied, diversity appeared in the identification of small-for GA neonates. These differences among countries disappeared when country specific reference centiles were utilized. The authors showed that the prevalence of small-for GA neonates correlated with the mean birthweight in each country, especially in very preterm infants (32).

In 2016, Greek national reference curves of fetal biometric parameters (biparietal diameter (BPD), occipitofrontal diameter, head circumference, femoral length (FL) and BPD/FL ratio) were published by Sotiriadis et al (33). These curves were based on 1200 singleton fetuses of Greek origin and they were compared with Intergrowth 21^st^ fetal and other previously published studies. The authors found statistically significant differences in certain parameters and they concluded that the use of other populations’ charts (e.g. Intergrowth 21^st^) may not be representative of national (local) population and may lead to improper classification of fetal growth status (33).

Even today, the choice of the appropriate centile charts is a matter of debate [34,35]. Kiserud et al. (6) commented on the proposed international use of normative WHO fetal growth charts that it is prudent to test whether the growth charts’ performance meets the local needs and that “*carefully adjusted growth charts reflecting optimal local growth should be used when public health issues are addressed*”. In the same line, the 2015 European Perinatal Health Report (35) commented that no consensus exists on the type (international versus national) of the most appropriate reference centile charts to be used and further research in Europe is imperative as growth restriction is an important perinatal health issue.

In the current study we developed and propose new population-based, sex-specific, birthweight for GA reference centiles for singleton neonates born in Greece. We also compared the centiles to other (inter) national population and Fenton centiles. The data provided by the Hellenic statistical authority derive from birth certificates which are mandatory in Greece; therefore, unregistered births almost do not exist. The vast majority of births take place in public or private obstetric clinics and birth related data are registered according to obstetricians’ or midwives’ records on individual births. Records are mostly complete as shown from the unavailability of birthweight, and/or GA in 1,92% of the neonates. Moreover, GA estimation, in nearly all pregnancies, is based not only on the date of the last menstrual period but also on an early ultrasound which is freely offered to pregnant women in Greece, minimizing the possibility of errors. In addition, the large sample size of this study that consisted of more than 600.000 neonates allowed for reasonable sized samples even in early GAs and made it feasible to perform statistical corrections for implausible birthweights at each GA.

The current study performed a comparison of the proposed reference centile chart with 9 other (inter)national centiles. The suggested centiles identified different small and large-for GA rates in comparison to the other centiles. For example, the proposed reference identified significantly less small-for GA neonates compared to the WHO and Intergrowth 21^st^ fetal centiles up to the 32^nd^ gestational week, an expected finding since the latter two are prescriptive fetal charts and the early GAs include infants that still stay in uterus. This similarity/diversity among the various centiles, may be attributed either to the study design (prescriptive versus descriptive-reference charts), the similarity/diversity of statistical methods employed (26) or it may be attributed to other factors such as ethnic variations (30,31) of the corresponding studied neonatal populations, maternal characteristics, different time periods of data collection, socioeconomic differences (36,37,38,39).

The definition of the last GA as 40+, due to the registration method of the Hellenic statistical authority, may be considered a study limitation. Due to this restriction, comparison at the 40^th^ week of GA was not attempted (Table 2). The cross-sectional nature of the current study may also be considered a limitation as no longitudinal data were collected or analyzed. Additional advanced statistical methods for the identification of implausible GAs exist. Such methods assume a mixture of more than two normal distributions of implausible neonatal birthweight within each early GA (40). We expect such methods to be very close to those applied and therefore, they would have a limited impact on the current study results. The present study may be included in future meta-analysis after systematic review. It may additionally be used for the development of local customized growth curves according to Gardosi’s approach. It may also be utilized for comparison with future Greek national curves and with national curves of other European countries, contributing to further research regarding the differences noticed in small and large-for GA neonatal rates in Europe (32,35).

## CONCLUSION

The proposed birthweight by gestational age reference centile curves are representative for the neonatal population born in Greece. They are based on contemporary cross-sectional data derived from a very large sample of over 600000 neonates born in 2011-2017. The centile curves could aid physicians to more accurately classify high risk neonates at birth and local stakeholders to plan public health research and perinatal economic health policies. Proper classification of neonates will also result in the appropriate early stage interventions with the prime aim to prevent short- and long-term adverse consequences.

### What is already known about the topic

➢ Three types of birthweight by GA charts are used for estimation of intrauterine growth, each created with different study design; prescriptive, descriptive and customized
➢ No consensus exists at the moment of the most appropriate type of chart to be used
➢ Contemporary birthweight by GA reference charts provide valuable information to neonatologists, public health researchers and policy makers

### What this study adds

➢ We propose contemporary, population based, nationally representative birthweight by GA reference charts, based on a large sample of over 600.000 singleton neonates born in Greece.
➢ The proposed charts can assist stakeholders in Greece to more accurately classify high risk infants, decrease adverse outcomes and efficiently allocate scarce perinatal healthcare resources

## Data Availability

The neonatal birth registry data are available from the Hellenic statistics authority (ELSTAT).

https://www.statistics.gr/en/statistics/-/publication/SPO03/-

## ACKNOWLEDGEMENTS

The birth data used were supplied by the Hellenic statistical authority (ELSTAT).

## DATA AVAILABILITY

The neonatal birth registry data are available from the Hellenic statistical authority (ELSTAT). (https://www.statistics.gr/en/statistics)

## AUTHOR CONTRIBUTIONS

AT and NV conceived the idea of the study. AT, KP and LD designed the study. KP and LD performed the statistical analysis. AT and KP drafted the manuscript and further revised after critical comments from NV, LD and EK. All authors have read and accepted the final manuscript.

