## Supplementary file for "Birthweight by gestational age reference centile charts for Greek neonates"

**Details of statistical methods for excluding outliers**

Firstly, a mixture of normal model was applied especially to early GA birthweight distributions. This mixture was applied in all GA distributions where the shape of the data implied that they were generated from at least two possibly overlapping normal distributions. In such cases, particularly for data between 22nd and 34th week, we fitted a mixture model of two normal distributions by the use of a maximum likelihood method via the use of an expectation-maximization algorithm (16,17). Relevant parameters were estimated, e.g. the mixing probabilities, the mean and standard deviations of each component distribution. In each GA we assumed two normal distributions with different means and standard deviations. The data were split into two groups based on their corresponding posterior probabilities. A probabilistic clustering of the observations was then obtained by allocating each observation to either group, according to the corresponding posterior probability of group membership.

Even though the expectation-maximization method could easily be applied when they are distinct systematic errors based on under-estimating GAs, in our study it did not perform properly when this distinction was not very clear; for example, in weeks 35-40. Nonetheless, even in these late GA strata, outliers do exist. The distributions in these GAs were normal, therefore an implausible birthweight was defined as being greater than 5 standard deviations away from the median birthweight for their GA and gender (18).

After removing outliers, each GA strata contained a minimum of 20 records of neonates and then exact percentiles of neonatal birthweights were calculated for each GA. Via univariate analysis we examined the birthweight distributions and additional neonatal characteristics.

**Figure A1. Birthweight (in grams) by gestational age (weeks) crude -not corrected- curves and centiles - Male neonates born between 2011 and 2017**

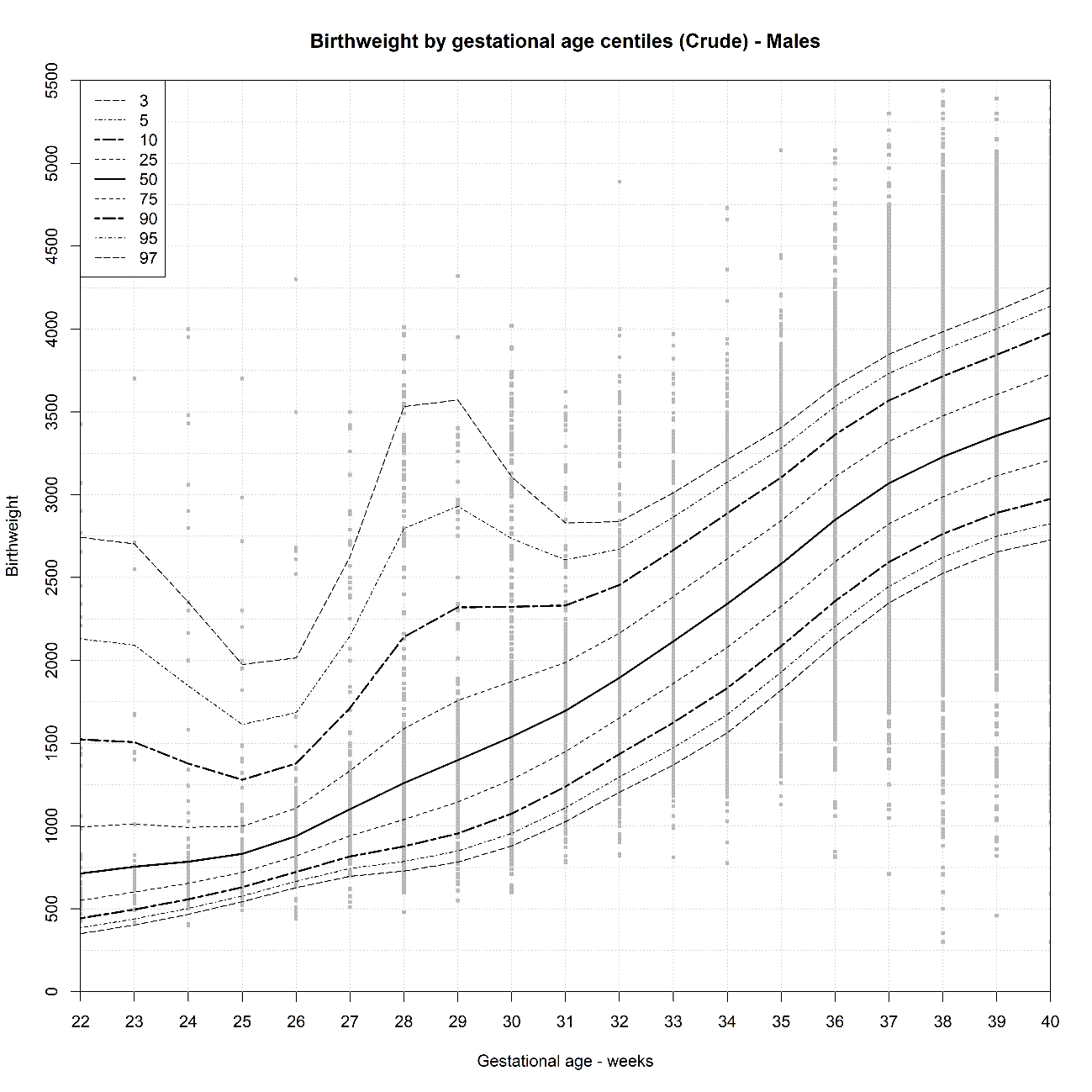

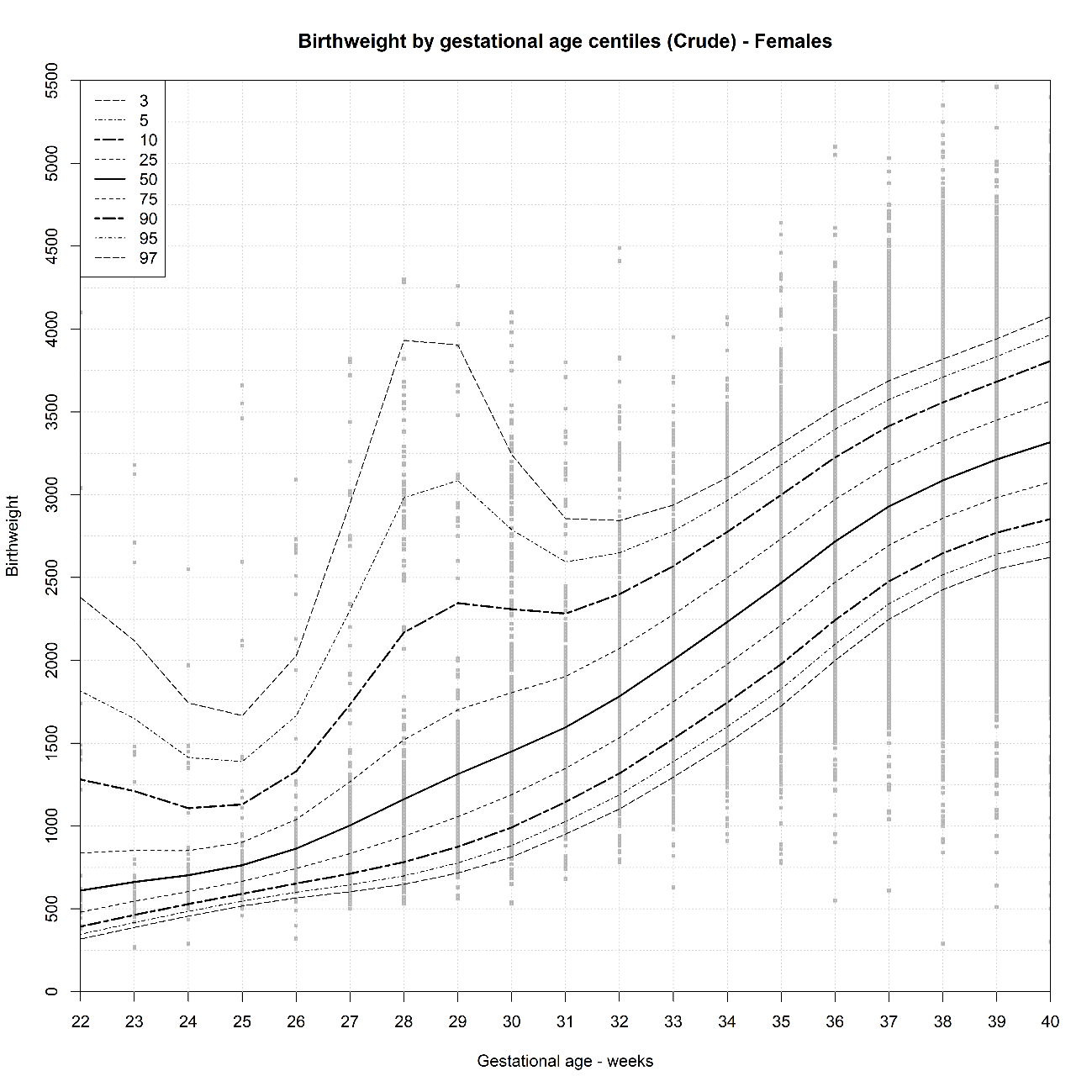
**Figure A2. Birthweight (in grams) by gestational age (weeks) crude -not corrected- curves and centiles Female neonates born between 2011 and 2017.**
